# Development of a Modular APEX2-based Lateral Flow Signaling Platform for Virus Diagnostics

**DOI:** 10.64898/2026.08.31.26361816

**Authors:** Noémi Kovács, Márk Kiss, Hajnalka Jankovics

## Abstract

Rapid point-of-care (POC) diagnostics require sensitive, stable, and cost-effective reporter systems. While lateral flow assays (LFAs) are widely used, their sensitivity is often limited by the use of colloidal gold, whereas enzymatic amplification using horseradish peroxidase (HRP) is constrained by the difficulties of recombinant production in bacterial hosts. In this study, we present a protein engineering framework utilizing APEX2, a robust engineered peroxidase, as a next-generation signaling element. We optimized the expression of APEX2-His6 in *E. coli*, achieving high yields (64 mg/L) of a functional apoenzyme that was efficiently reconstituted *in vitro* (RZ value: 2.1). Furthermore, we investigated the development of a bifunctional reporter by genetically fusing APEX2 to a single-domain antibody (sdAb). While the production of the fusion protein encountered significant solubility challenges in *E. coli* (yielding ∼0.5 µg/liter culture), our results confirm that APEX2 retains high catalytic activity and stability when integrated into an LFA format. Using West Nile Virus (WNV) as a model system, we demonstrate that APEX2-mediated enzymatic amplification provides a clear colorimetric signal on nitrocellulose membranes. This work establishes APEX2 as a commercially viable, recombinantly accessible alternative to traditional peroxidases, providing a versatile platform for the development of high-sensitivity diagnostic tools.

**Graphical Abstract:** 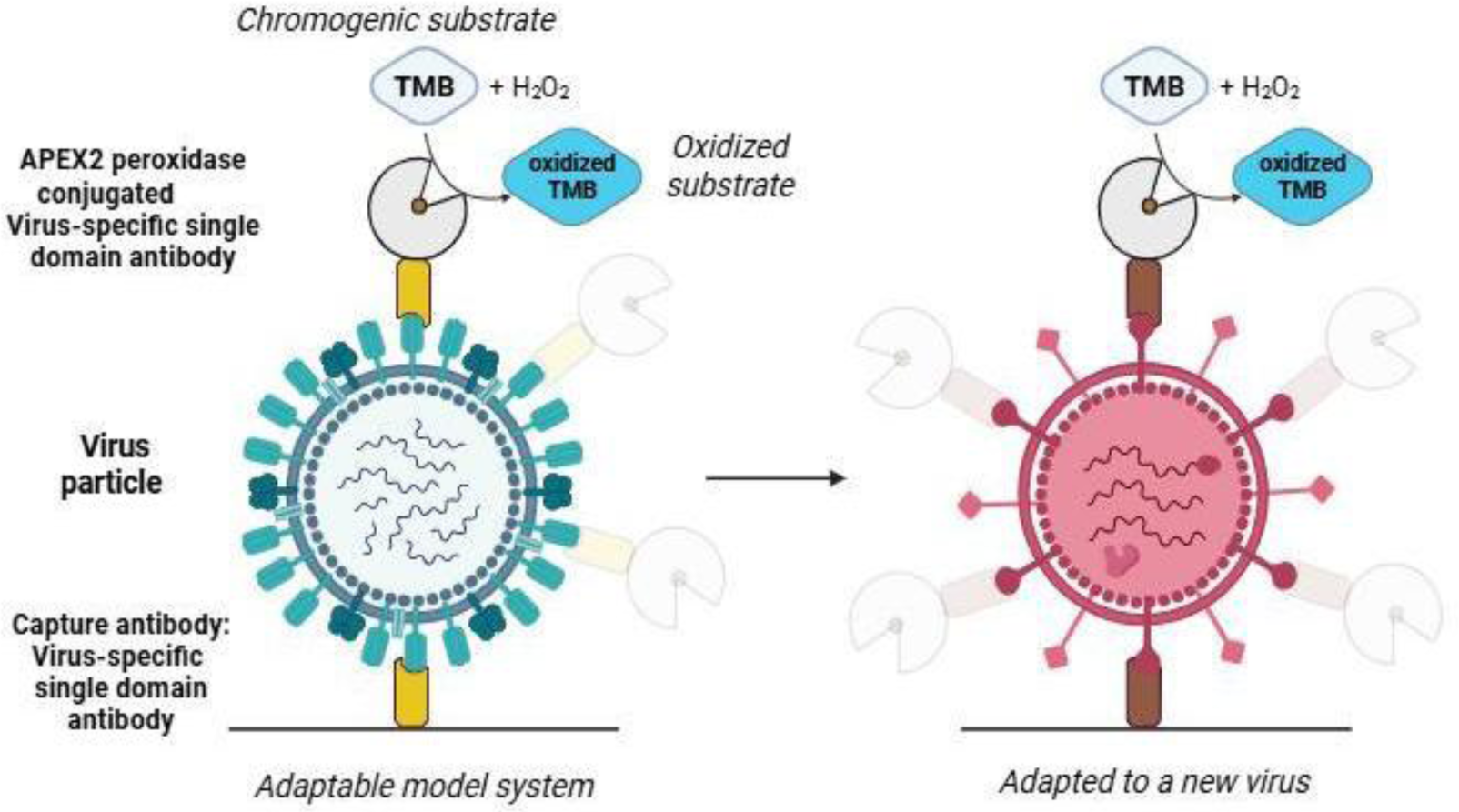

*Created in* https://BioRender.com

## Introduction

The rapid and accurate detection of viral pathogens is a cornerstone of global public health, especially in the face of emerging viruses and variants driven by human-induced environmental changes and high viral mutation rates (Cassedy et al., 2021; Ryu, 2017). Effective diagnostic tools are essential for clinical management and the prevention of widespread outbreaks. Currently, the diagnostic landscape is divided between nucleic acid-based methods and immunoassays (Lin et al., 2020). While techniques like PCR and RT-PCR offer high sensitivity and specificity, they require specialized equipment and significant time, which limits their use in resource-limited or field settings.

Immunoassays, such as ELISA and lateral flow assays (LFAs), provide a more accessible alternative (Bahadır & Sezgintürk, 2016). LFAs, in particular, are favored for point-of-care (POC) testing due to their speed and ease of use. However, conventional LFAs often rely on colloidal gold nanoparticles, which may lack the sensitivity required to detect low analyte concentrations in early-stage infections (Mulvaney et al., 2020). Integrating enzymatic signal amplification into the LFA platform can address this limitation, yet the production of the most common reporter, horseradish peroxidase (HRP), remains problematic.

Traditional HRP-based assays are hindered by the enzyme’s complex folding requirements, including essential disulfide bonds and post-translational glycosylations, which make its recombinant expression in bacterial systems inefficient (Hwang & Espenshade, 2016; Lee et al., 2015). To overcome these challenges, APEX2, a genetically engineered variant of soybean ascorbate peroxidase, has been developed as a robust alternative. APEX2 is a monomeric, non-glycosylated protein that can be efficiently expressed in the cytoplasm of *E. coli* while maintaining superior stability and catalytic potency (Lam et al., 2014; Lee et al., 2015; Sherwood & Hayhurst, 2019).

The performance of such diagnostic platforms also depends on the recognition elements used. Conventional IgG antibodies face limitations due to their large size and the need for complex production systems (Belfakir et al., 2025; Jin et al., 2023). Single-domain antibodies (sdAbs or nanobodies) offer a compelling solution (Zhang et al., 2025). Due to their small size, high thermal stability, and ability to recognize cryptic epitopes, sdAbs are ideal for biosensor applications (Muyldermans, 2013). Furthermore, their single-gene structure allows for the creation of genetic fusions with reporter enzymes, simplifying the manufacturing process.

In this study, we present a modular diagnostic platform based on the genetic fusion of an anti-WNV nanobody and the APEX2 peroxidase. We utilize West Nile Virus (WNV) as a model system to validate the platform. WNV is a mosquito-borne flavivirus that represents an increasing public health threat in both North America and Europe (Gupta et al., 2026; Hamer et al., 2008; Paz, 2015). By optimizing the production and reconstitution of APEX2, we aim to establish a framework for next-generation, enzymatically amplified LFAs that combine the robustness of nanobodies with the high sensitivity of engineered peroxidases.

## Results

### Design and Architecture of the sdAb-APEX2-Based Lateral Flow Platform

To achieve rapid and sensitive detection of viral antigens, we designed a sandwich-type lateral flow assay (LFA) that integrates the high specificity of nanobodies with the potent enzymatic signal amplification of APEX2 peroxidase. The architecture of the system is based on three modular, recombinantly produced protein components, allowing for easy adaptation to emerging pathogens. The functional layout and the step-by-step mechanism of the assay are illustrated in **Figure 1**.

**Figure 1.**
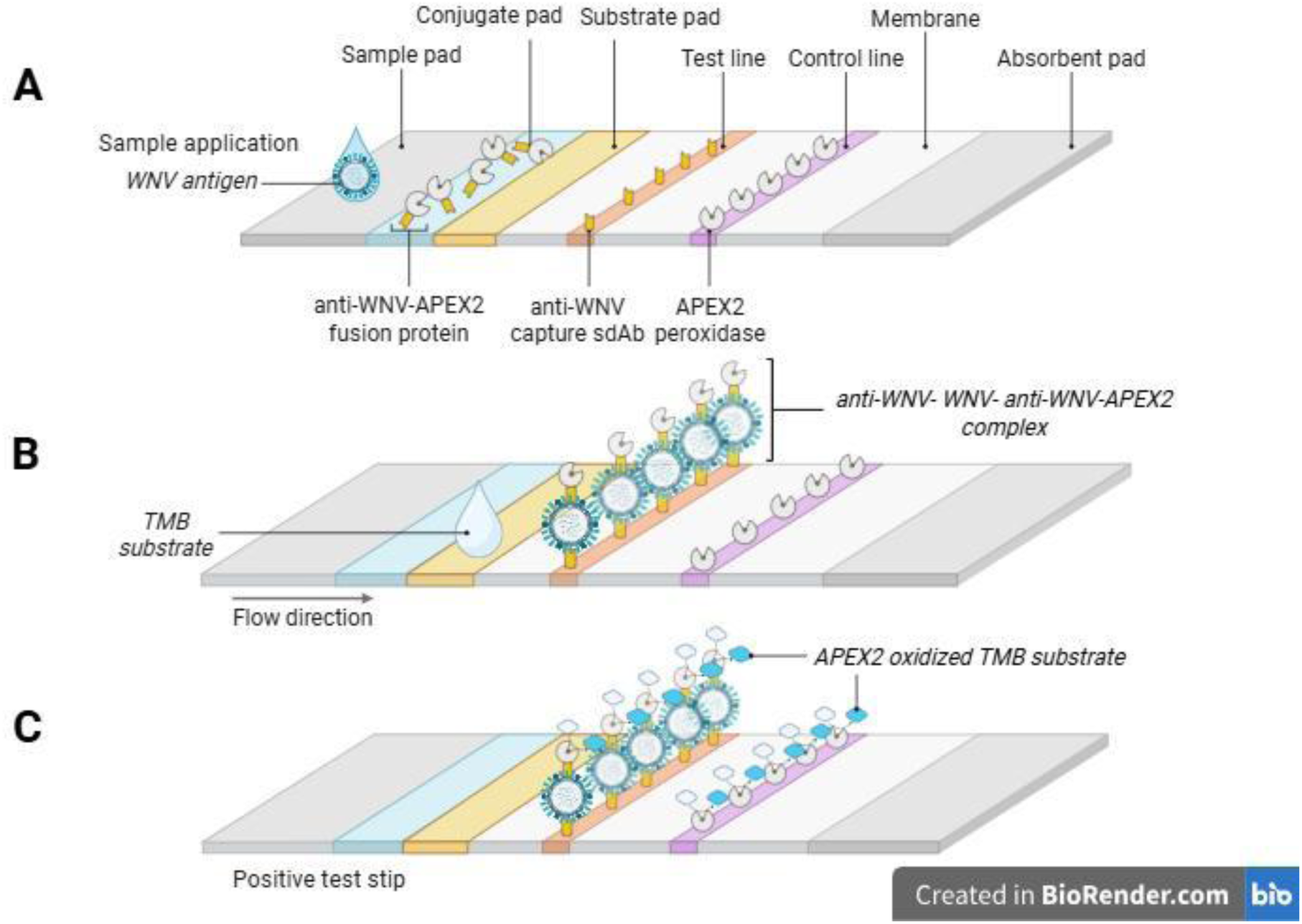
Schematic architecture and operational principle of the sdAb-APEX2-based lateral flow platform. (A) Target recognition and complex formation: Upon sample application, WNV antigens are captured by the bifunctional sdAb-APEX2 detection conjugate on the conjugate pad, forming a mobile immune complex. **(B) Sandwich assembly and substrate initiation:** The complex is immobilized by capturing antibodies at the Test line. Following capillary migration, the TMB chromogen substrate is applied directly to the membrane to initiate enzymatic signaling. **(C) Visual readout and functional validation:** APEX2-mediated oxidation of TMB results in a localized blue color at the Test line in positive samples. The Control line, containing immobilized APEX2-His6, generates a blue signal regardless of the antigen presence, confirming the catalytic activity of the system and proper reagent flow.

The assay relies on the coordinated action of the following components and steps:

1. **Bifunctional Detection Conjugate (sdAb-APEX2):** The core of the signaling system is a fusion protein consisting of an anti-WNV single-domain antibody (sdAb_A10_) and the APEX2 peroxidase enzyme. As shown in **Figure 1A**, this conjugate is pre-deposited on the conjugate pad. We selected the sdAbA10 nanobody due to its exceptional affinity and specificity for domain III (DIII) of the WNV envelope glycoprotein (Hruškovicová et al., 2022). Upon sample application, this bifunctional reagent specifically captures the WNV antigens, forming a mobile immune complex.
2. **Capture Zone and Substrate Application:** The nitrocellulose membrane is functionalized with an immobilized anti-WNV capture antibody at the Test line. As the sample fluid migrates through the membrane via capillary action, this antibody “sandwiches” the antigen–sdAb–APEX2 complex at the specific site. To visualize the accumulated enzyme, the TMB (3,3’,5,5’-tetramethylbenzidine) chromogen substrate is applied directly to the membrane after the migration is complete (**Figure 1B**).
3. **Visual Readout and Functional Control:** The final assay result is determined by the localized enzymatic oxidation of the substrate. The APEX2 enzyme, concentrated at the Test line by the captured antigen, catalyzes the conversion of TMB into a visible blue product (**Figure 1C**). To validate the assay’s performance, a Control line is established using immobilized APEX2-His6 enzyme. This line serves as an internal functional check: it reacts with the added TMB regardless of the presence of the antigen, producing a second blue band that confirms both the successful flow of reagents and the catalytic activity of the peroxidase system (**Figure 1C**).

The use of an *E. coli*-based expression system for all protein components ensures cost-effective and scalable production, making this platform a versatile model for rapid diagnostic development where high sensitivity is achieved through direct enzymatic signal generation.

### Production and functional characterization of the recombinant APEX2-His6 peroxidase

To provide a robust signaling component and reliable control for the LFA platform, we optimized the production of the APEX2-His6 peroxidase in *E. coli BL21(DE3)pLysS* cells. Initially, we attempted to generate the active holoenzyme *in vivo* by supplementing the culture medium with 25 µM hemin to facilitate cofactor uptake into the periplasm. During this optimization, we found that hemin preparation was critical; the cofactor was only effective when dissolved in a DMSO/NaOH-based solution, as it exhibited poor solubility in ammonium acetate buffer alone. However, upon scaling up production, this *in vivo* approach encountered significant technical limitations: high concentrations of residual hemin in the cell lysate interfered with the immobilized metal affinity chromatography (IMAC) purification by displacing Ni²⁺ ions from the chelating resin. To overcome this bottleneck, we refined the protocol to express and purify APEX2 as an apoenzyme (**Figure 2**). This strategy allowed for efficient IMAC purification, yielding up to 64 mg of high-purity protein per liter of culture, which was subsequently reconstituted into the active holoenzyme after dialysis.

**Figure 2.**
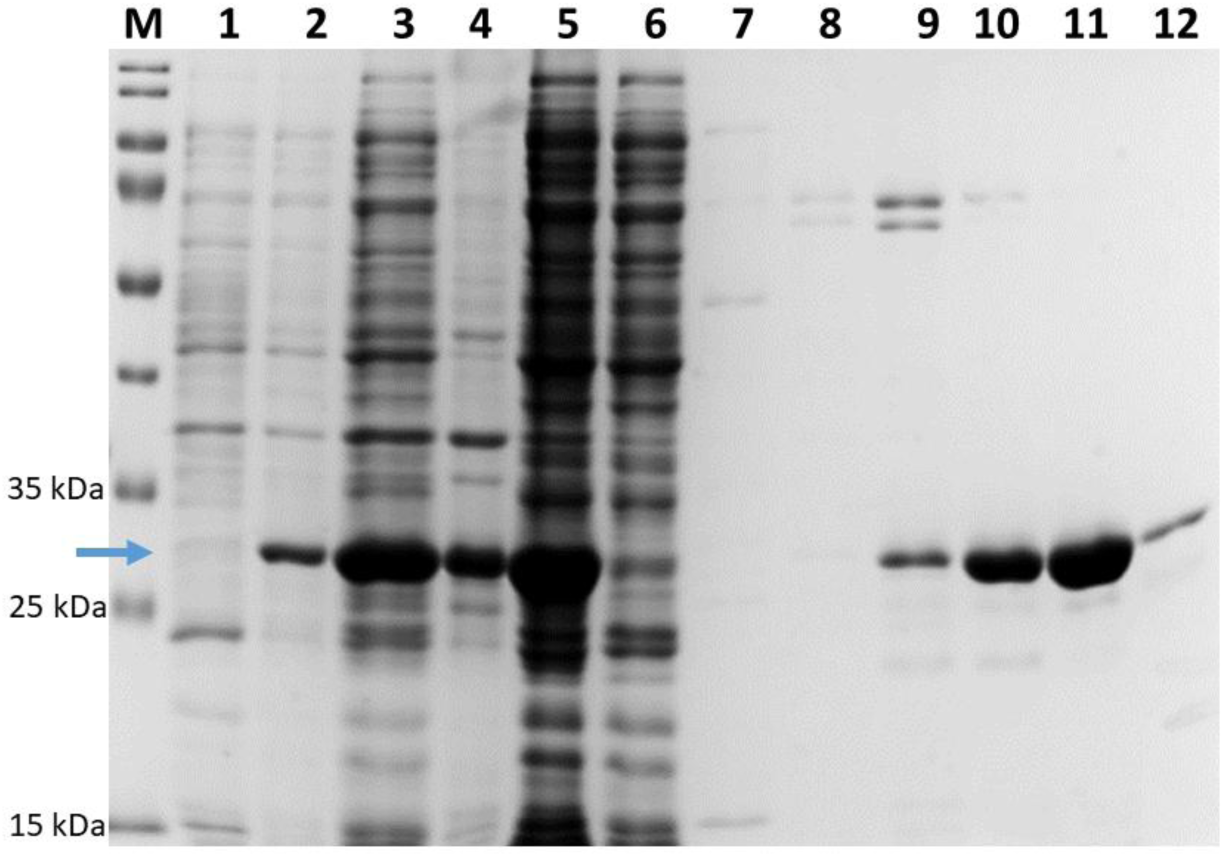
SDS-PAGE analysis of the expression and purification of APEX2-His6. The blue arrow indicates the target protein at the expected molecular weight of 28.15 kDa. **Lane M:** PageRuler Prestained Protein Ladder; **Lanes 1–2:** Total cell lysates before and after induction with IPTG; **Lanes 3–5:** Comparison of the cell suspension, pellet, and the filtered soluble cell extract; **Lane 6:** Flow-through fraction; **Lane 7:** Column wash; **Lanes 8–12:** Purified fractions of the elution peak showing high homogeneity of the recombinant peroxidase. ract, (Lane 6) Flow- through, (Lane 7) Second column wash, (Lanes 8–12) Fractions of the elution peak.

The transition from the inactive apoenzyme to the catalytically active holoenzyme was achieved through a controlled *in vitro* reconstitution process with hemin. Successful heme incorporation and proper folding were confirmed by UV-Vis spectrophotometry, where the reconstituted holoenzyme exhibited a characteristic Soret band with an absorbance peak at 403 nm. The calculated RZ value (Reinheitszahl, A_403_/A_280_) reached approximately 2.1, indicating a high degree of heme saturation and exceptional protein purity. To verify that the recombinant APEX2 maintains its enzymatic potency, we performed functional ELISA-like assays. The results demonstrated that the reconstituted APEX2-His6 effectively catalyzes the oxidation of the TMB substrate, producing a robust and stable colorimetric signal. This high catalytic activity confirms that the *E. coli*-derived APEX2 is fully functional and suitable for both the signaling conjugate and the internal control line of the LFA system.

### Challenges in the expression and solubility of the sdAb-APEX2 bifunctional conjugate

While the production of the standalone APEX2-His6 was highly efficient, the development of the genetic fusion with the anti-WNV sdAb_A10_ nanobody presented significant biotechnological challenges. Initial expression trials using the pET23b(+) vector in both *E. coli BL21(DE3)pLysS* and *SHuffle T7 Express* strains failed to yield reproducible protein bands on SDS-PAGE, suggesting that the basal expression of the fusion construct might be toxic or lead to rapid degradation.

To achieve controlled expression, the genetic construct was transferred to the pET28a(+) vector, which utilizes the more strictly regulated T7lac promoter. This switch successfully induced high-level protein production; however, the resulting sdAb-APEX2 fusion protein was primarily localized in inclusion bodies as an insoluble aggregate. To enhance the solubility and proper folding of the bifunctional conjugate, we incorporated a flexible (Gly4Ser)_2_ linker and an enterokinase (EK) cleavage site between the domains (**Figure 3**).

**Figure 3.**
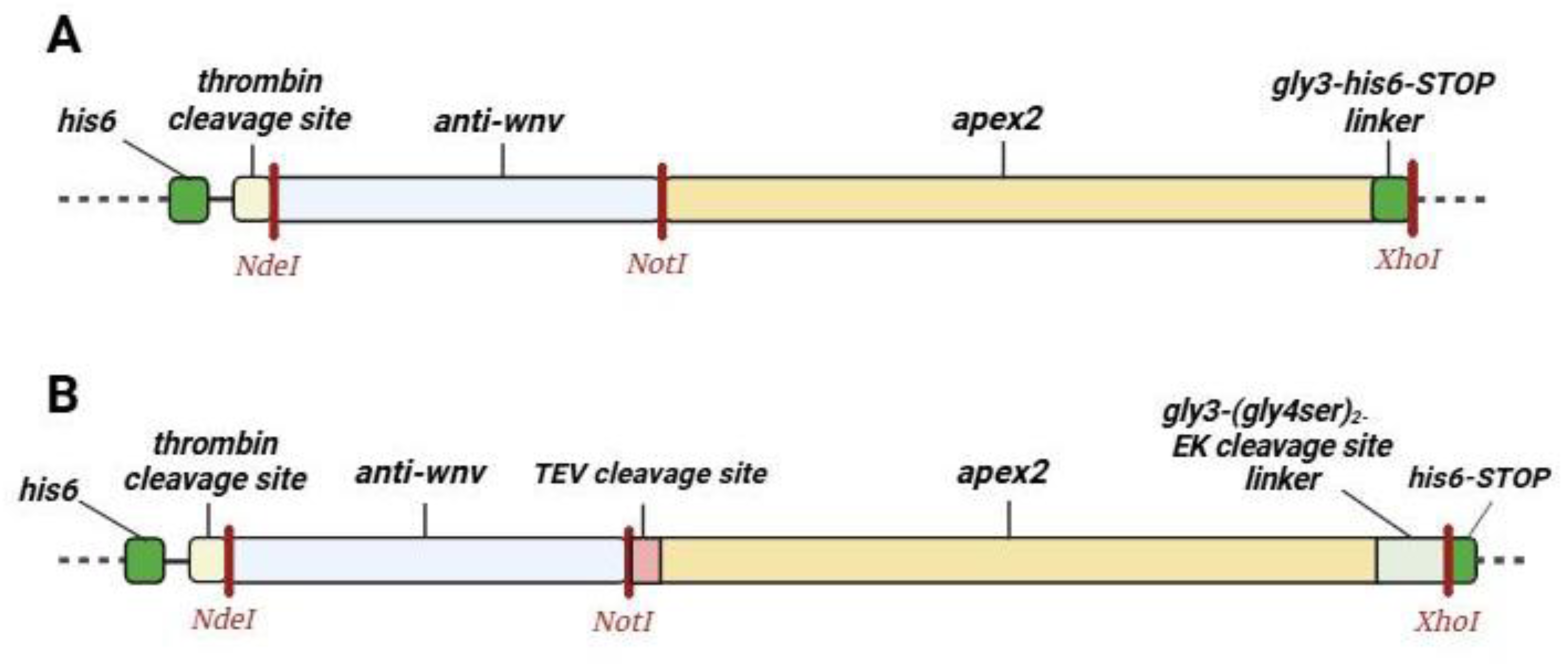
Schematic representation of the genetic constructs for the sdAb-APEX2 bifunctional conjugate. **(A)** Initial design of the anti-WNV-APEX2-His6 fusion protein. **(B)** Optimized expression cassette in the pET28a(+) vector, incorporating a flexible (Gly4Ser)_2_ linker and an enterokinase (EK) cleavage site between the anti-WNV sdAb and the APEX2 peroxidase to improve folding and solubility. The T7lac promoter was utilized to ensure strictly regulated expression. Created in https://BioRender.com

The anti-WNV sdAbA10 nanobody was also attempted to be produced independently; however, these attempts did not yield the desired results, as the protein could not be expressed in a soluble form at sufficient quantities. To address this, we incorporated a TEV cleavage site between the nanobody and the APEX2 peroxidase in our fusion construct. This allows for the potential extraction of the sdAb if the fusion protein can be successfully expressed.

Furthermore, we systematically optimized the induction parameters, reducing the temperature to 16 °C and the IPTG concentration to 50 µM.

Under these refined conditions in the *E. coli SHuffle* strain, we successfully achieved the expression of the sdAb-APEX2 conjugate in a soluble form. However, the yield remained significantly lower compared to the standalone peroxidase, reaching only the submicrogram- per-culture range (approx. 0.5 mg/ liter culture). While this confirms the feasibility of producing the bifunctional reporter, the current yields are insufficient for a full-scale characterization and direct application on LFA test strips. These results highlight a critical bottleneck in the production of nanobody-peroxidase fusions: while both components are highly stable individually, their genetic fusion appears to interfere with the folding kinetics or the solubility of the complex in *E. coli*.

Consequently, to demonstrate the operational principle of our LFA platform in this study, we utilized the high-yield APEX2-His6 as a model enzyme, while the optimization of the bifunctional conjugate remains a target for future protein engineering efforts (e.g., through chaperone co-expression or alternative secretion pathways).

## Discussion

In recent decades, human-induced environmental degradation and the rapid rate of viral mutation have led to the emergence of numerous pathogenic viral strains and variants. This challenge is further exacerbated by the faster spread of pathogens driven by globalization and urbanization, as well as the expanding geographic range of viral vectors due to global warming. Highly pathogenic pathogens - such as COVID-19 - can trigger pandemics and cause severe public health and economic damage; consequently, rapid and easily adaptable detection methods are needed to prevent their spread. Immunodiagnostic platforms, such as ELISA or LFA, are ideally suited for this purpose. At the same time, it is essential to overcome the bottlenecks that hinder or slow down the development of these tests.

The transition from conventional antibodies to alternative protein scaffolds in point-of-care diagnostics is already reflected in commercial applications. A prominent example is the cPass™ SARS-CoV-2 neutralization test (GenScript), which utilizes the recombinant ACE2 receptor instead of antibodies to functionally detect neutralizing responses (Tan et al., 2020). Similarly, nanobodies comparable in sensitivity to LFA tests using conventional antibodies have been exploited in lateral flow assays for diagnostics (Bates et al., 2026).

While conventional lateral flow assays (LFAs) typically exhibit lower sensitivity compared to ELISA - often by orders of magnitude - the integration of enzymatic amplification can bridge this diagnostic gap. As summarized by (Gong et al., 2025), enzyme-linked signal enhancement on nitrocellulose membranes can improve the limit of detection (LOD) by up to 100-fold. Our APEX2-sdAb construct leverages this principle; by utilizing the high catalytic activity of APEX2, we provide a streamlined pathway to high-sensitivity point-of-care diagnostics.

Our APEX2-sdAb fusion construct follows this technological trajectory, combining the high affinity of single-domain antibodies with the direct enzymatic signaling of an engineered peroxidase in a single, robust polypeptide chain.

The production of conventional immunoglobulins, which are commonly used in immunoassays, requires complex and time-consuming mammalian expression systems. Single-domain antibodies (sdAbs) with high binding affinity offer an attractive alternative to conventional IgG antibodies, as they can be produced more quickly and easily in bacterial expression systems. Another area of development is the replacement of horseradish peroxidase (HRP) as a reporter molecule. Due to HRP’s complex, glycosylated structure - which contains multiple disulfide bonds as well as heme and calcium cofactors - it is primarily isolated from horseradish in its native form and then retroactively conjugated to IgG molecules via chemical conjugation. The efficiency of this reaction significantly influences the results, often leading to nonspecific HRP binding or steric hindrance at the antibody binding sites. To overcome these limitations, APEX2 - a genetically engineered soybean L-ascorbate peroxidase - was developed. APEX2 is free of glycosylation and disulfide bonds, functions effectively even under reductive conditions, requires only a heme cofactor, and can be produced quickly and easily in *E. coli* expression systems. Furthermore, APEX2 can be expressed as a direct fusion protein with sdAbs, thereby avoiding the negative consequences of conjugation and ensuring a defined 1:1 antibody–peroxidase stoichiometry.

Considering these factors, we developed an sdAb-APEX2-based protein model system that integrates these technological advancements and can be easily adapted for the detection of emerging viruses and variants in ELISA and LFA formats. Using West Nile virus (WNV) as a model, we demonstrated the functional characteristics of the three essential protein components, their bacterial production in *E. coli*, and their purification via immobilized metal affinity chromatography (IMAC). To ensure the quality control of the study, we optimized the high-yield production of APEX2 peroxidase as an apoenzyme in *E. coli BL21(DE3)pLysS* cells, competent for apoenzyme production, and subsequently reconstructed it into an active holoenzyme using hemin. For specific virus binding, we selected the sdAb_A10_ antibody (anti-WNV) directed against domain III of the WNV envelope glycoprotein and engineered the anti-WNV-APEX2 fusion protein to detect the target molecule. Due to the poor reproducibility of antibody production and its tendency to aggregate, we optimized the structural composition and expression parameters of the fusion protein using the *E. coli SHuffle* strain and successfully achieved partial solubility.

To improve purification efficiency and solubility, we incorporated a linker containing a (Gly4Ser)₂-EK cleavage site into the C-terminal end of the sdAb-APEX2 construct, immediately upstream of the H6 hexahistidine tag. In addition, the insertion of a TEV protease cleavage site between the anti-WNV and APEX2 domains allows for the recovery of the functional antibody from the fusion protein via controlled proteolytic digestion. Overall, this strategy demonstrates that fusing difficult-to-express, aggregation-prone antibodies to APEX2 offers a viable approach for their production and functional integration.

## Methods

### Construction of the plasmids coding the proteins

For the design of the proteins and, subsequently, the expression plasmids, we used the sequences published earlier. The gene of the anti-WNV was taken from the FR1-FR4 VHH region of the sdAb_A10_ antibody (Hruškovicová et al., 2022). We designed APEX2 without the N-terminal Flag tag and with a C-terminal tag consisting of a Gly3-His6 coding sequence (Sherwood & Hayhurst, 2019). The sequences were codon-optimized for *E. coli*; the genes were synthesized by Twist Bioscience (San Francisco, California, Canada) and inserted into the pET28a(+) plasmid between the NdeI and XhoI restriction sites, respectively, resulting in the first version of the anti-WNV-APEX2-His fusion protein.

For special purposes the fusion protein is completed with a TEV protease cleavage site and a C-terminal linker peptide. In that construct, we used PCR reactions with Q5 polymerase to incorporate the TEV protease cleavage site into the N-terminus of APEX2, while at the C-terminus, following Gly3, we inserted the (Gly4Ser)₂-Enterokinase protease cleavage site (EK) linker sequence. We digested the modified APEX2 gene and the original pET28a(+) -*anti-wnv- apex2-his6* plasmid using Not-HF and XhoI enzymes. The digested vector and the insert was purified from the agarose gel using the Monarch DNA Gel Extraction Kit (New England Biolabs, Ipswich, Massachusetts, USA), then ligated with T4 ligase (Thermo Scientific Waltham, Massachusetts, USA). *E. coli* TOP10 competent cells (Agilent Technologies, Santa Clara, California, USA) were transformed with the ligation mixture, and the cells were plated onto LB agar plates containing 50 µg/mL kanamycin. The pET28b(+)-*anti-wnv-tev-apex2- gly3-(gly4ser)_2_-ek-his6* plasmid DNA was purified from selected colonies, then analyzed by restriction digestion and sequencing. DNA sequencing was performed by Macrogen Europe (Amsterdam, the Netherlands).

For the production of the APEX2-His6 protein, its coding sequence was inserted between the NotI and XhoI restriction sites of the pET23b(+) plasmid.

### Expression and Purification of APEX2-His6

The APEX2-His6 gene was expressed in *E. coli BL21(DE3)pLysS* cells using a pET23b(+) vector. Cells were cultured in LB medium supplemented with 100 µg/mL ampicillin and 20 µg/mL chloramphenicol at 37 °C until an OD_600_ of 0.6–1.0 was reached.

In initial optimization experiments, the medium was supplemented with 25 µM hemin (prepared as a 5 mM stock in a DMSO / 0.1 N NaOH / 1.4 M ammonium acetate buffer) to promote *in vivo* holoenzyme formation. However, due to hemin-mediated displacement of Ni^2+^ ions during subsequent purification, the protocol was modified to produce the apoenzyme. Expression was induced with 0.1 mM IPTG at 25 °C for 3 hours.

Cells were harvested by centrifugation and resuspended in Buffer A (20 mM NaH_2_PO_4_, 500 mM NaCl, 25 mM imidazole, pH 7.5) with EDTA-free protease inhibitor (Roche, Basel, Switzerland). After disruption by sonication, the lysate was clarified by two-step centrifugation (10,000xg followed by 84,000xg) and filtered through a 0.45µm PES membrane. The protein was purified using a 5mL HiTrap HP Chelating column (Ni−NTA) on an ÄKTA Start FPLC system (GE Healthcare). After washing with 25mM imidazole, the APEX2−His6 apoenzyme was eluted with Buffer B (containing 500 mM imidazole). Purified fractions were analyzed by SDS−PAGE and dialyzed against 10mM NaH_2_PO_4_, 150 mM NaCl (pH 7.4), followed by concentration measurements at A_280_ using a molar extinction coefficient of 14,440 M^-1^cm^-1^.

### Reconstitution and Soret-band Measurement

The purified APEX2 apoenzyme was converted into the active holoenzyme by adding hemin (2.5 mM hemin dissolved in 0.1 N NaOH) in a 1:1 molar ratio. The mixture was incubated for 30 minutes at room temperature. To determine the success of cofactor incorporation and protein purity, UV-Vis spectra were recorded in the 250–500 nm range using a spectrophotometer and quartz cuvettes. The Reinheitszahl (RZ) value was calculated as the ratio of the Soret band absorbance (at 403 nm) to the protein absorbance (at 280 nm).

### Enzymatic Activity Assay (ELISA-based)

The peroxidase activity of the reconstituted APEX2-His6 was validated using a modified ELISA protocol. Recombinant APEX2 was immobilized on high-binding microtiter plates. After blocking with 5% BSA, TMB (3,3’,5,5’-tetramethylbenzidine) substrate was added. The reaction was monitored by the development of a blue color, and the enzymatic activity was quantified by measuring the absorbance after stopping the reaction with 1 M H_2_SO_4_.

### Production Challenges of anti-WNV and anti-WNV-APEX2-His6 fusion protein

*SHuffle T7 Express* competent *E. coli* cells were transformed by the optimized pET28(+)*-anti-wnv-tev-apex2- his6* plasmid, and were spread onto LB-Agar plates containing 50 µg/ml kanamycin, then grown overnight at 30°C.

The fusion protein was expressed by the inoculation of a 7 mL LB medium containing 50 µg/ml kanamycin and grown overnight at 30 °C (260 rpm). 500 mL fresh TB/Kan medium was supplemented with 1% (v/v) overnight culture and grown at 30 °C until OD_600_ reached 1. Cell culture was induced by 50 μM IPTG and further incubated at 16 °C, overnight with shaking. Cells were harvested by centrifugation at 4,000xg (40 min at 6.0 °C) using a Heraeus Biofuge Multifuge centrifuge (Hanau, Germany) and subsequently kept at -20 °C.

Cells in 10 mL buffer ‘A’ (20 mM NaH_2_PO_4_, 500 mM NaCl, 25 mM imidazole, pH 8.0) supplemented with EDTA-free protease inhibitor (Roche, Basel, Switzerland) and incubated on ice for an hour. The cells were disrupted by sonication (Cole-Parmer, Vernon Hills, Illinois, USA). The remaining cell debris was partitioned by centrifugation (using 10,000×g on an Heraeus Biofuge primo R for 30 min at 6°C) and the supernatant was centrifuged again at 84,000×g on an Optima Max-XP Ultracentrifuge (Beckman Coulter, Brea, California, USA) for 30 min at 10 °C The supernatant filtered through a 0.45 µm pore size PES syringe filter.

A 5 mL HisTrap FF Chelating Ni-affinity column connected to an ÄKTA Start FPLC system (GE Healthcare) was equilibrated with buffer ‘A’. After loading the filtrate, the column was washed with 35 ml buffer ‘A’, 35 mL buffer ‘A’ containing 25 mM imidazole, and 35 mL buffer ‘A’ containing 75 mM imidazole. The protein was eluted in one step with a 35 ml buffer ‘B’ containing 500 mM imidazole. Protein samples were analyzed by SDS-PAGE, then the fractions containing the purified fractions were combined, followed by concentration measurements at A_280_ using a molar extinction coefficient of 48,485 M^-1^cm^-1^.

## Data Availability

All data produced in the present study are available upon reasonable request to the authors.

## Acknowledgement

Project no. 2025-2.1.1-EKÖP-2025-00029/92 has been implemented with the support provided by the Ministry of Culture and Innovation of Hungary from the National Research, Development and Innovation Fund, financed under the 2025-2.1.1-EKÖP funding scheme.

